# The Role of Oestrogen in Female Skeletal Muscle Ageing: A Systematic Review

**DOI:** 10.1101/2023.05.18.23290199

**Authors:** Annabel J. Critchlow, Danielle Hiam, Ross Williams, David Scott, Séverine Lamon

**Affiliations:** School of Exercise and Nutrition Sciences, Institute for Physical Activity and Nutrition (IPAN), Deakin University, Geelong, Australia

**Author notes:** Corresponding author: A/Prof. Séverine Lamon, School of Exercise and Nutrition Sciences, Faculty of Health, Deakin University, 221 Burwood Hwy, Burwood 3125., Australia, Ph.

**Keywords:** ageing, ovarian hormones, menopause, skeletal muscle.

## Abstract

Ageing is associated with a loss of skeletal muscle mass and function that negatively impacts the independence and quality of life of older individuals. Females demonstrate a distinct pattern of muscle ageing compared to males, potentially due to menopause where endogenous sex hormone production declines. This systematic review aims to investigate the current knowledge about the role of oestrogen in female skeletal muscle ageing. A systematic search of MEDLINE complete, Global Health, Embase, PubMed, SPORTDiscus, and CINHAL was conducted. Studies were considered eligible if they compared a state of oestrogen deficiency (e.g. postmenopausal females) or supplementation (e.g. oestrogen replacement therapy) to normal oestrogen conditions (e.g. premenopausal females or no supplementation). Outcome variables of interest included measures of skeletal muscle mass, function, damage/repair, and energy metabolism. Quality assessment was completed with the relevant Johanna Briggs critical appraisal tool, and data were synthesised in a narrative manner. Thirty-two studies were included in the review. Compared to premenopausal females, postmenopausal females display reduced muscle mass and strength, but the effect of menopause on markers of muscle damage and expression of the genes involved in metabolic signalling pathways remains unclear. Some studies suggest a beneficial effect of oestrogen replacement therapy on muscle size and strength, but evidence is largely conflicting and inconclusive, potentially due to large variations in the reporting and status of exposure and outcomes. The findings from this review points toward a potential negative effect of oestrogen deficiency in ageing skeletal muscle, but further mechanistic evidence is needed to clarify its role.

**Graphical abstract figure:** 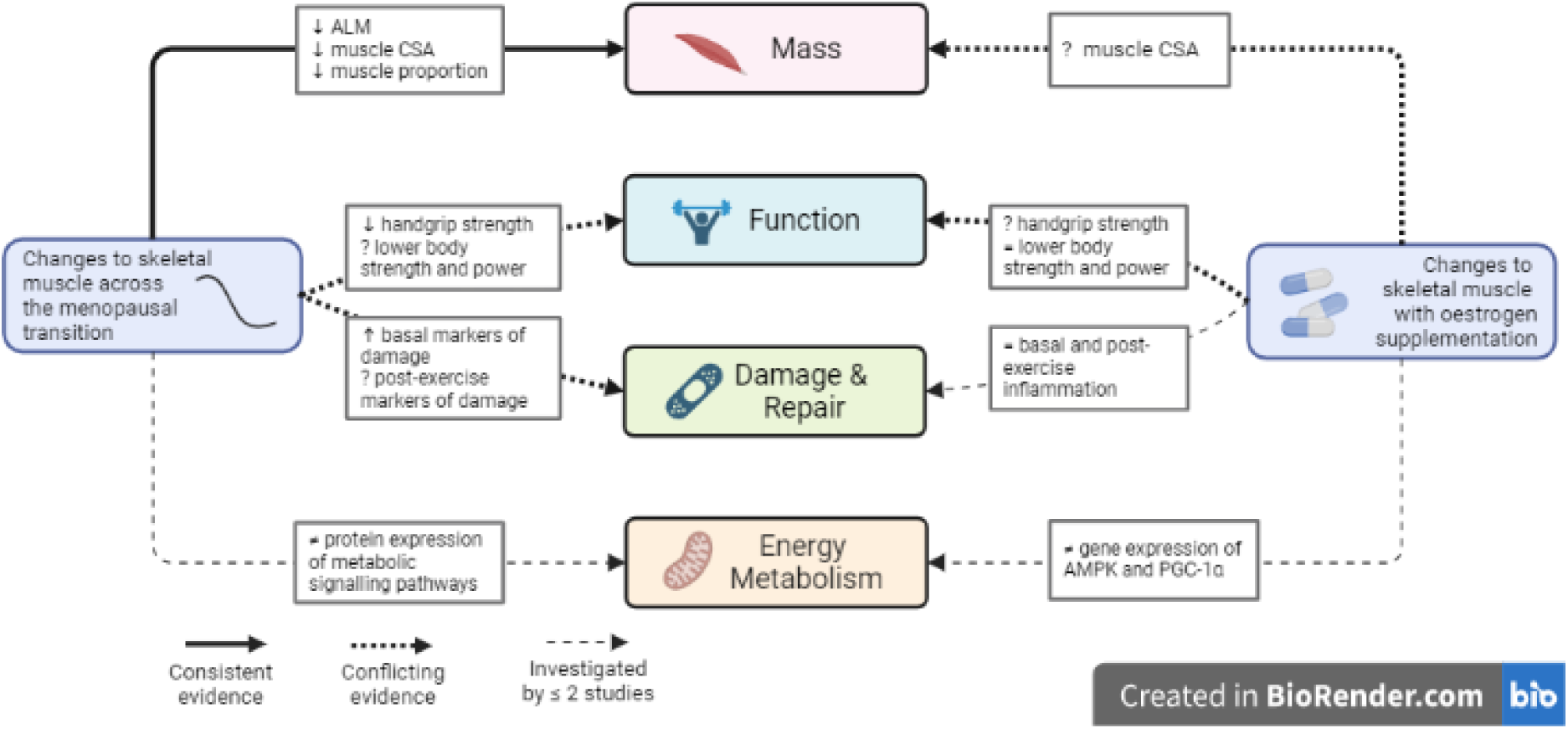

The role of oestrogen in female skeletal muscle ageing. ↑ = significant increase, ↓ = significant decrease, ≠ = significantly different, ? = mixed evidence, p<0.05. ALM: appendicular lean mass; AMPK: adenosine monophosphate kinase; CSA: cross-sectional area; PGC-1α: peroxisome proliferator-activated receptor gamma coactivator 1-alpha. Created with BioRender.com.

## Introduction

Skeletal muscle ageing is associated with an insidious and progressive loss of muscle mass concomitant to a reduction in muscle strength (Cruz-Jentoft & Sayer, 2019). Independent of disease, muscle mass and strength decline by approximately 8% and 15% per decade from the fourth decade of age, with a rapid acceleration after 70 years (Kim & Choi, 2013). This reduction in skeletal muscle health directly contributes to impaired physical function, impacting the independence and quality of life of older adults who may become unable to carry out daily living activities (Visser *et al*., 2005). Amongst the cellular impairments responsible for this age-induced reduction in muscle mass and function, older individuals demonstrate a blunted anabolic response to exercise or nutritional stimuli compared to young individuals. This is further compounded by increased chronic inflammation, mitochondrial dysfunction, impaired autophagy, a reduction in the number and myogenic potential of satellite cells and a decrease in the number of motor units, leading to muscle fibre atrophy and impaired force production (reviewed in Wiedmer *et al*. (2021)).

Most of the evidence regarding the mechanisms of muscle ageing originates from male human or animal models. Skeletal muscle structure and function is however highly sex-specific, with ∼3,000 genes being differentially expressed between the sexes (Oliva *et al*., 2020). Generalising male findings to female populations without consideration of the sex-specific context of skeletal muscle ageing is hence inappropriate. Females typically live longer than males but have worse physical disability (Murtagh & Hubert, 2004), a higher prevalence of low muscle mass (Janssen *et al*., 2002) and experience a faster loss of muscle strength in the fifth decade of life (Haynes *et al*., 2020). On the molecular level, De Jong *et al*. (2023) recently demonstrated that, while the molecular processes altered by ageing in male and female muscle are similar, the magnitude of change to these pathways varies between the sexes.

Sex differences in skeletal muscle remodelling during ageing partly arise from disparities in the sex hormone profile. In males, positive relationships exist between testosterone, the major androgen hormone, and muscle mass and function over the lifespan (Bhasin *et al*., 2001; Mouser *et al*., 2016). In females, three forms of oestrogen, the major ovarian hormone, are produced endogenously: oestrone (E1), oestradiol (E2), and oestriol (E3) (Cui *et al*., 2013). Although their main role is ascribed to reproduction, oestrogens also target non-reproductive tissues, including skeletal muscle (Wiik *et al*., 2009), where they bind to oestrogen receptor α (ERα) and β (ERβ) to regulate the transcription of their target genes (Heldring *et al*., 2007). Levels of oestrogen vary throughout the female lifespan and are specific to each of the bioactive forms. E2 fluctuates across the menstrual cycle and across the female lifespan, where its levels rise at the onset of puberty, peak between 15 and 20 years, gradually decline during the transition to menopause and remain low until death. On the other hand, E1 becomes the predominant oestrogen post-menopause. Finally, a large spike in E3 is observed during pregnancy due to its production in the placenta (reviewed in Cui *et al*. (2013)).

Numerous studies have attempted to correlate fluctuations in ovarian hormones with muscle mass and strength in pre- and postmenopausal females, but to which extent oestrogen, or a lack thereof, regulates muscle health during ageing is unclear. This systematic review summarises the literature investigating the role of oestrogen on female skeletal muscle health during ageing by focusing on its effects on muscle mass, composition and function, and on molecular measures of skeletal muscle health including regeneration, inflammation, mitochondrial function and substrate metabolism.

## Methods

This systematic review was conducted and reported in accordance with the Preferred Reporting Items for Systematic Reviews and Meta-Analyses (PRISMA 2020, see Supplementary File 1 and 2) and was registered in the international prospective register of systematic reviews (PROSPERO) (CRD42022374366). The Assessing the Methodological Quality of Systematic Reviews 2 (AMSTAR2) tool was used to appraise the systematic review (Supplementary File 3). Figure 1 depicts the search process. The complete protocol, including database searching, eligibility criteria, screening, quality assessment, data extraction and synthesis can be found in Supplementary File 4.

**Figure 1:**
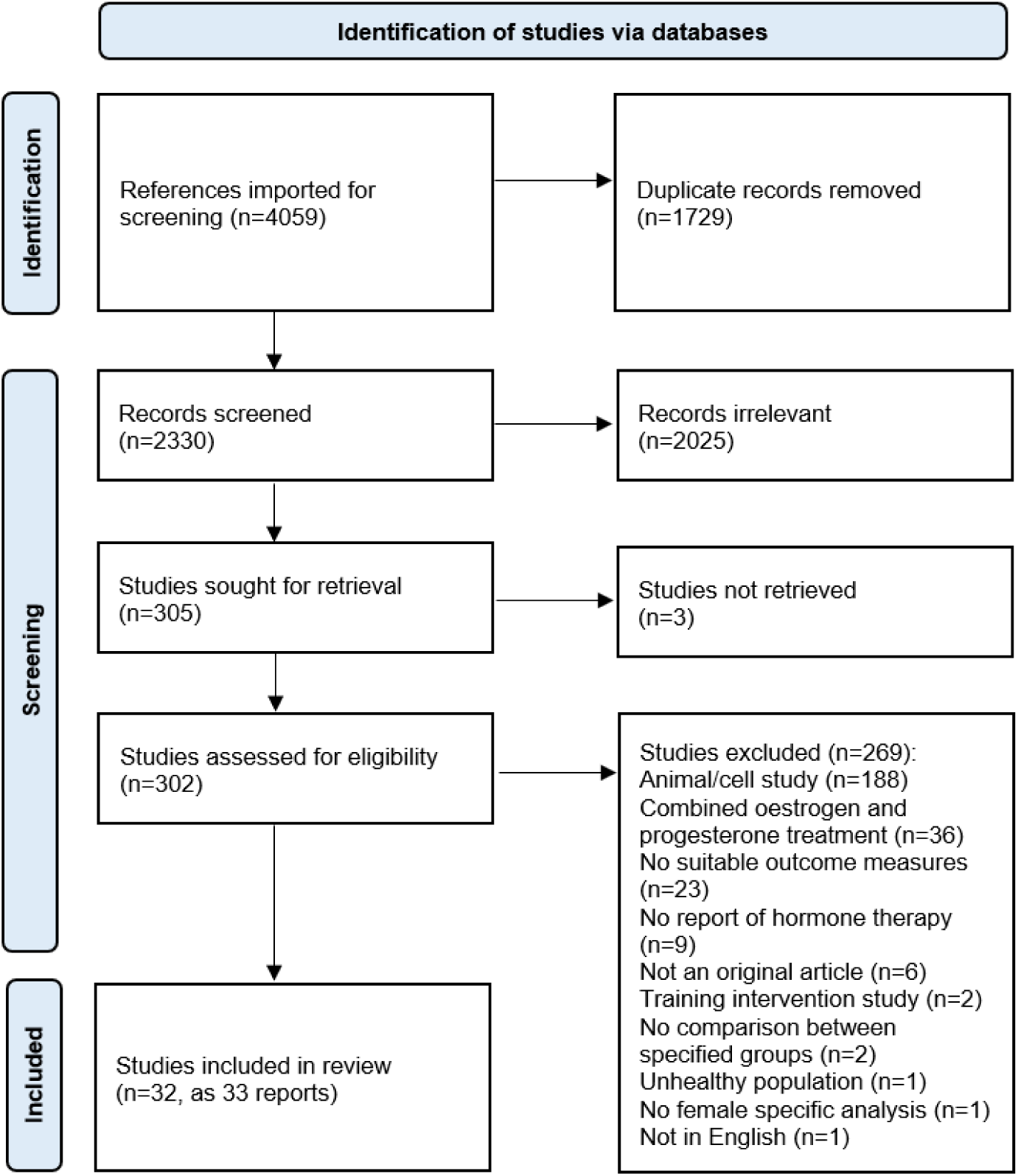
PRISMA flow diagram of the systematic search and assessment.

## Findings

### Quality assessment

Following the search process described in Figure 1, thirty-two studies were assessed according to their study design (cross-sectional, quasi-experimental, randomised controlled trial (RCT) or longitudinal). Overall, 19 studies (59%) had a low risk, 10 studies (31%) had a moderate risk, and three studies had a high risk (9%) of bias. Full details of the quality assessment can be found in Supplementary File 5.

### Associations between oestrogen and skeletal muscle health in postmenopausal females

#### Muscle mass

Seven studies ranging from 49 to 1244 participants investigated the associations between endogenous oestrogen and skeletal muscle mass, determined by appendicular lean mass (ALM), in postmenopausal females (Table 1). There was no association between bioavailable (Gonnelli *et al*., 2014) or total serum E2 levels (Kenny *et al*., 2003; Rolland *et al*., 2007; Kong *et al*., 2019; Guligowska *et al*., 2021) and ALM or the loss of ALM with age. There was a potential, positive correlation between serum E2 levels and ALM index (ALMI; ALM normalised to height) when ALMI was derived from bioimpedance analysis (BIA) (Guligowska *et al*., 2021), but not when ALMI was derived from DXA (García-Martín *et al*., 2013; Gonnelli *et al*., 2014).

**Table 1:** Studies investigating associations between oestrogen levels and skeletal muscle outcomes.

| Study | Study Design | Participants (mean age ± SD) | Independent variable | Dependent variable | Associations |
| --- | --- | --- | --- | --- | --- |
| *Bochud et al. (2019)* | Cross-sectional | 366 pre- and postmenopausal females between 18-90 years old ($52 \pm 16$ years) | Urinary E1<br>Urinary E2 | Handgrip strength<br>Handgrip strength | No association<br>No association |
| *Garcia-Martin et al. (2013)* | Cross-sectional | 91 postmenopausal females >45 years old ($56 \pm 4$ years) | Serum E2 | ALMI | No association |
| *Gonelli et al. (2014)* | Cross-sectional | 854 postmenopausal females >55 years old ($64 \pm 6$ years) | Free E2 index | ALM<br>ALMI | No association<br>No association |
| *Guligowska et al. (2021)* | Cross-sectional | 61 elderly community dwelling females >75 years (median: 79 years) | Serum E2 | ALM<br>ALMI<br>Handgrip strength | No association<br>Positive association ($rS=0.289$)<br>No association |
| *Kenny et al. (2003)* | Cross-sectional | 189 postmenopausal females >59 years old using ORT for >2 years ($68 \pm 5$ years) | Serum E2 | ALM | No association |
| *Kong et al. (2019)* | Cross-sectional | 1244 postmenopausal community dwelling females 40-69 years old ($71 \pm 7$ years) | Serum E2 | ALM<br>Handgrip strength | No differences between E2 tertiles |
| *Rolland et al. (2007)* | Longitudinal (Screened at baseline, 6, 12, 24, and 36 months) | 49 postmenopausal females ($54 \pm 4$ years) | Serum E1<br>Serum E2 | Loss of ALM<br>Loss of muscle strength<br>Loss of ALM<br>Loss of muscle strength | No association<br>Positive association ($r=0.29$)<br>No association<br>No association |

ALM: appendicular lean mass; ALMI: appendicular lean mass index; ORT: oestrogen replacement therapy; RCT: randomised controlled trial; RM: repetition maximum.

#### Muscle function

Four of these studies also investigated the associations between endogenous oestrogen and muscle function (Table 1). Serum E2 and urinary E1 and E2 levels were not correlated with handgrip strength (Bochud *et al*., 2019; Kong *et al*., 2019; Guligowska *et al*., 2021). Higher serum E1, but not E2, was associated with greater loss of knee extensor strength over 3 years in 49 postmenopausal females (Rolland *et al*., 2007).

Overall, there was no or inconclusive associations between oestrogen levels and skeletal muscle mass and function in postmenopausal females.

### Effect of menopausal status on skeletal muscle health

#### Muscle mass

Table 2 outlines the key findings of the 17 studies that investigated differences in skeletal muscle outcomes depending on menopausal status. Muscle mass-related outcome measures were investigated in ten of these studies. ALM, ALMI and leg lean mass were significantly lower in postmenopausal versus pre- and peri-menopausal females in most (Nyberg *et al*., 2017; Juppi *et al*., 2020; Park *et al*., 2020; Rathnayake *et al*., 2021), but not all studies (Smith *et al*., 2014). Similarly, thigh muscle cross-sectional area (CSA) was significantly smaller in post-compared to premenopausal females in some (Pöllänen *et al*., 2015; Juppi *et al*., 2020), but not all studies (Pöllänen *et al*., 2011; Ahtiainen *et al*., 2012), and it was decreased in a small study comparing 14 late- to 13 early-postmenopausal females (Park *et al*., 2019). Relative thigh muscle proportion was also lower in post-versus pre-menopausal females in one study (Pöllänen *et al*., 2011), but not another (Ahtiainen *et al*., 2012). Differences in the composition and molecular regulation of muscle mass were also investigated. Myofiber CSA was not different in the *vastus lateralis* of peri-versus postmenopausal females (Collins *et al*., 2019). No differences in fibre type proportion were found in the *vastus lateralis* muscle, yet muscle gene expression of anabolic signalling molecules insulin-like growth factor (IGF) isoforms 1Ea and 1Ec were lower in a study comparing 15 post- to 14 premenopausal females (Ahtiainen *et al*., 2012).

**Table 2:** Studies investigating the effect of menopausal status on skeletal muscle outcomes.

| Study | Participants (mean age ± SD) | Intervention | Determination of Menopausal Status | Relevant Outcome Measures | Key Findings |
| --- | --- | --- | --- | --- | --- |
| *Ahtiainen et al. (2012)* | 14 premenopausal females ($32 \pm 2$ years)<br>15 postmenopausal monozygotic twin females ($58 \pm 2$ years) | N/A | Unknown | Thigh muscle CSA, *vastus lateralis* gene expression of IL-6 and IGF | ↔ relative thigh muscle area (premenopausal: $54.4 \pm 11.1\%$, postmenopausal: $49.8 \pm 12.7\%$)<br>↔ muscle IL-6<br>↓ muscle IGF-1Ea and IGF-1Ec levels in postmenopausal females (72% and 60% of premenopausal values, respectively)<br>↔ muscle IGF-1R, IGFBP3, and IGFBP5 levels |
| *Arnett et al. (2000)* | 15 premenopausal females ($23 \pm 7$ years)<br>10 postmenopausal females ($59 \pm 11$ years) | Exercise session to induce muscle damage: participants performed 6 sets of 10 reps of eccentric hamstring curls at 110% of their concentric 1-RM | Unknown | Serum CK | ↑ baseline serum CK in postmenopausal ($102.0 \pm 38.5$ U/L) versus premenopausal ($32.0 \pm 14$U/L) females<br>↓ post-exercise serum CK in postmenopausal versus premenopausal females at 72 and 96hr<br>Inverse association between serum E2 and resting serum CK ($r=-0.38$, 14% of variation in serum CK explained by serum E2) |
| *Bassey et al. (1996)* | 89 regular premenopausal females ($48 \pm 3$ years)<br>33 females with irregular menstrual periods (unspecified menopausal stage, $50 \pm 3$ years)<br>92 postmenopausal females ($51 \pm 3$ years) | N/A | Unknown | Handgrip and isometric quadriceps strength, and leg extensor power | ↔ handgrip strength (regular: $31 \pm 7$N, irregular: $31 \pm 5$N, postmenopausal: $29 \pm 5$N)<br>↔ isometric quadriceps strength (regular: $399 \pm 86$N, irregular: $409 \pm 99$N, postmenopausal: $380 \pm 127$N)<br>↔ leg extensor power (regular: $171 \pm 60$W, irregular: $181 \pm 61$W, postmenopausal: $163 \pm 56$W) |
| *Buckley-Bleiler et al. (1989)* | 8 younger premenopausal females ($24.6 \pm 3.5$ years)<br>10 older premenopausal females ($43.6 \pm 2.2$ years)<br>6 postmenopausal females ($52.8 \pm 2.1$ years) | Exercise session to induce muscle damage: participants completed 40 maximal effort isometric muscle actions of right thigh, held for 10s with 20s rest | Young premenopausal: regular menstrual cycles of $\sim 31$ days<br>Old premenopausal: regular menstrual cycles of $\sim 27$ days<br>Postmenopausal: cessation of menstrual periods for $>2$ years<br>serum E2 $<130$pmol.L-1 | Isometric MVC and serum CK | ↔ pre-exercise MVC (younger: $378 \pm 23$N, older: $366 \pm 17$N, postmenopausal: $342 \pm 25$N)<br>↔ strength loss during exercise<br>↔ pre-exercise serum CK (younger: $31 \pm 3$IU/L, older: $30 \pm 3$IU/L, postmenopausal: $34 \pm 6$IU/L)<br>↔ post-exercise serum CK at 6, 24, 48, and 72 hours |
| *Collins et al. (2019)* | 5 perimenopausal females ($53 \pm 1$ years)<br>5 postmenopausal females ($54 \pm 1$ years) | <1 year follow up | Assessed by STRAW criteria and unspecified serum FSH and E2 levels | *Vastus lateralis* myofibre CSA and satellite cell number | ↔ myofibre CSA<br>↔ satellite cell number<br>Positive association between serum E2 and satellite cell number ($r^2=0.478$) |
| *Juppi et al. (2020)* | 89 early perimenopausal females ($51 \pm 2$ years)<br>145 late perimenopausal females ($52 \pm 2$ years)<br>197 postmenopausal females followed up from baseline ($53 \pm 2$ years) | Participant visits every 3-6 months to determine menopausal status until they become postmenopausal | Perimenopausal: unspecified FSH levels and menstrual diary<br>Postmenopausal: FSH >30IU/mL and no menstrual bleeding in previous 5-6 months | ALM, ALMI, lean mass of leg, absolute and relative thigh muscle area, *vastus lateralis* myofibre type distribution and size | ↔ ALM, ALMI, leg lean mass, absolute and relative thigh muscle area between early and late perimenopausal<br>↓ ALM ($17.8 \pm 2.2$kg versus $18.0 \pm 2.2$kg), ALMI ($6.5 \pm 0.6$kg/m<sup>2</sup> versus $6.6 \pm 0.6$kg/m<sup>2</sup>), and leg lean mass ($6.7 \pm 0.8$kg versus $6.8 \pm 0.9$kg) in postmenopausal versus perimenopausal females<br>↓ absolute ($165.3 \pm 10.1$ cm<sup>2</sup> versus $166.9 \pm 9.6$cm<sup>2</sup>) and relative ($68.9 \pm 6.0\%$ versus $69.6 \pm 5.6\%$) thigh muscle area in postmenopausal versus perimenopausal females<br>↔ myosin isoform proportion of type I, IIA and IIX fibres |
| *Laakkonen et al. (2017)* | 6 premenopausal females ($32 \pm 2$ years)<br>9 postmenopausal monozygotic twin females ($57 \pm 2$ years) | N/A | Unknown | Handgrip strength, lower-limb muscle power (countermovement jump), and specific force | ↓ handgrip strength in postmenopausal females ($258.9 \pm 36.1$N versus $333.6 \pm 45.7$N)<br>↓ lower-limb muscle power in postmenopausal females ($12.6 \pm 3.8$cm versus $26.6 \pm 5.7$cm)<br>↔ specific muscle force (postmenopausal: $9.8 \pm 1.7$N/cm<sup>2</sup>, premenopausal: $10.1 \pm 1.8$N/cm<sup>2</sup>)<br>Differentially expressed proteins involved in mitochondrial dysfunction, oxidative phosphorylation, glycolysis, TCA cycle, tyrosine degradation, pyruvate formation, glycogen biosynthesis, and ILK signalling |
| *Nyberg et al. (2017)* | 20 premenopausal females ($50 \pm 0$ years)<br>16 early postmenopausal females ($54 \pm 1$ years) | Aerobic exercise training: 12 weeks of high intensity training on a cycle ergometer for 1 hour, 3 times a week | Premenopausal: unspecified menstrual cycle regularity<br>Postmenopausal: no menstrual bleeding for >12 months and <5 years, with unspecified serum FSH and LH levels | Leg lean mass, venous-arterial lactate difference, and protein content of mitochondrial complexes (I to V) | ↓ baseline leg lean mass in postmenopausal females ($13.1 \pm 0.4$kg versus $14.3 \pm 0.4$kg)<br>↔ baseline venous-arterial lactate difference<br>↔ baseline protein content of mitochondrial complexes (I to V) |
| *Park et al. (2019)* | 13 early postmenopausal females ($\leq 6$ years post-menopause, $55 \pm 3$ years)<br>14 late postmenopausal females ($\geq 10$ years post-menopause, $62 \pm 3$ years) | N/A | Unknown | Thigh muscle CSA | ↓ thigh muscle CSA in late postmenopausal females ($97.1 \pm 15.9$cm<sup>2</sup> versus $107.8 \pm 11.1$cm<sup>2</sup>) |
| *Park et al. (2020)* | 30 premenopausal females ($38 \pm 6$ years)<br>31 early perimenopausal females ($50 \pm 3$ years)<br>30 late perimenopausal females ($50 \pm 4$ years)<br>26 early postmenopausal females ($55 \pm 3$ years)<br>27 late postmenopausal females ($62 \pm 4$ years) | N/A | STRAW criteria | ALM and ALMI | ↓ ALM with progressing menopausal status (premenopausal: $17.8 \pm 1.7$kg, early perimenopausal: $18.7 \pm 2.7$kg, late perimenopausal: $16.8 \pm 2.7$kg, early postmenopausal: $17.6 \pm 3.1$kg, late postmenopausal: $16.0 \pm 2.6$kg)<br>↓ ALMI in late perimenopausal and late postmenopausal compared to early perimenopausal<br>↓ ALMI in prior HRT users in early and late postmenopausal stages versus non-users<br>No association between serum E2 and ALMI |
| *Pesonen et al. (2021)* | 25 early perimenopausal females ($51 \pm 2$ years)<br>38 late perimenopausal females ($52 \pm 2$ years) | N/A | STRAW criteria,<br>Early perimenopausal: FSH <25IU/L<br>Late perimenopausal: FSH between 25-30IU/L | Maximal isometric knee extension strength, dorsiflexor and plantarflexor MVC, twitch force potentiation, and vertical jump height | ↓ MVC plantarflexion in late ($108.8 \pm 32.4$Nm) versus early ($131.1 \pm 33.9$Nm) perimenopausal females<br>↔ MVC dorsiflexion (early: $31.9 \pm 9.1$Nm, late: $34.3 \pm 6.7$Nm)<br>↔ vertical jump height (early: $0.19 \pm 0.05$m, late: $0.20 \pm 0.04$m)<br>↔ knee extensor strength (early: $162.2 \pm 27.9$Nm, late: $153.7 \pm 38.8$Nm)<br>↔ twitch force potentiation (early: $10.1 \pm 6.3\%$, late: $6.7 \pm 9.3\%$) |
| *Pollanen et al. (2011)* | 13 premenopausal females ($33 \pm 2$ years)<br>13 postmenopausal females ($64 \pm 2$ years) | N/A | Unknown | Quadriceps femoris muscle CSA, relative thigh muscle area, quadriceps attenuation, knee extensor strength, and muscle force per CSA | ↔ quadriceps muscle CSA (pre: $58 \pm 8$cm<sup>2</sup>, post: $50 \pm 8$cm<sup>2</sup>)<br>↓ relative muscle area within muscle compartment in postmenopausal females ($94 \pm 2\%$ versus $95 \pm 3\%$)<br>↓ knee extension in postmenopausal females ($386 \pm 58$N versus $514 \pm 115$N)<br>↓ muscle force per CSA in postmenopausal females ($8.3 \pm 0.9$N/cm<sup>2</sup> versus $9.9 \pm 1.6$N/cm<sup>2</sup>)<br>↓ quadriceps attenuation in postmenopausal females ($55 \pm 4$HU versus $61 \pm 4$HU)<br>Positive correlation between serum E2 and muscle force per area ($r=0.411$) and quadriceps attenuation ($r=0.674$)<br>Positive association between serum E1 and muscle force per area ($r=0.596$) and quadriceps attenuation ($r=0.535$)<br>Inverse association between intramuscular E2 and quadriceps attenuation ($r=-0.469$)<br>No association between intramuscular E2 and muscle force per area |
| *Pollanen et al. (2015)* | 8 premenopausal females (32 ± 2 years)<br>8 postmenopausal females (58 ± 2 years) | N/A | Unknown | Thigh CSA, knee extensor strength, vertical jump height | ↓ thigh CSA in postmenopausal females (92.2 ± 12.2cm<sup>2</sup> versus 106.1 ± 11.4cm<sup>2</sup>)<br>↓ knee extensor strength in postmenopausal females (437.7 ± 90.6N versus 584.2 ± 106.8N)<br>↓ vertical jump height in postmenopausal females (13.5 ± 4.4cm versus 26.4 ± 6.1cm) |
| *Preisinger et al. (1995)* | 18 premenopausal females (22 ± 0 years)<br>22 postmenopausal females (60 ± 2 years) | N/A | Unknown | Handgrip strength | ↓ dominant handgrip strength in postmenopausal females (155 ± 8.3N versus 208 ± 10.5N)<br>↓ non-dominant handgrip strength in postmenopausal females (136.9 ± 6.7N versus 180.7 ± 9.6N) |
| *Rathnayake et al. (2021)* | 184 premenopausal females (42 ± 6 years)<br>166 postmenopausal females (56 ± 4 years) | N/A | STRAW criteria | ALM, relative muscle mass, and handgrip strength | ↓ ALM in postmenopausal females (14.8 ± 2.9kg versus 16.0 ± 2.5kg)<br>↓ ALMI in postmenopausal females (6.6 ± 1.0kg/m<sup>2</sup> versus 6.9 ± 0.9kg/m<sup>2</sup>)<br>↓ Handgrip strength in postmenopausal females (15.2 ± 4.8kg versus 19.0 ± 6.0kg)<br>Positive association between serum E2 and grip strength in postmenopausal (r=0.16) but not premenopausal females |
| *Romero-Parra et al. (2021)* | 19 premenopausal females (29 ± 6 years)<br>13 postmenopausal females (52 ± 4 years) | Exercise session to induce muscle damage: 10 sets of 10 reps of back squats (4 seconds in eccentric phase, 1 second pause, and 1 second concentric phase) at 60% of 1-RM, with 2 minutes of rest between sets | Unknown | Back squat 1RM, 1RM per kg/FFM, countermovement jump, myoglobin, muscle soreness, serum LDH and CK | ↓ 1-RM in postmenopausal females (50.7 ± 4.2kg versus 74.6 ± 17.3kg)<br>↓ 1-RM/kg FFM in postmenopausal females (1.3 ± 0.2kg versus 1.8 ± 0.4kg)<br>↑ baseline serum myoglobin in postmenopausal females (76.9 ± 13.8µg/L) compared to all 3 menstrual cycle phases (early follicular: 62.2 ± 13.4µg/L, late follicular: 60.4 ± 7.5µg/L, mid-luteal: 60.1 ± 10.9µg/L)<br>↔ baseline and peak serum CK, LDH, countermovement jump, muscle soreness |
| *Smith et al. (2014)* | 12 premenopausal females (33 ± 2 years)<br>24 postmenopausal females (61 ± 2 years) | N/A | Unknown | ALM, basal protein synthesis rate, mRNA expression of MyoD, myostatin, follistatin, and FOXO3 | ↔ ALM between pre- (19.4 ± 1.0kg) and postmenopausal (17.8 ± 0.5kg) females<br>↑ basal protein synthesis rate in post- versus pre-menopausal females (+20%)<br>↑ MyoD, follistatin, and FOXO3 mRNA in post- versus premenopausal females (+40-90%)<br>↔ myostatin between pre- and postmenopausal females |

↑ = significant increase, ↓ = significant decrease, ↔ no difference, $p < 0.05$ . ALM: appendicular lean mass; ALMI: appendicular lean mass index; CK: creatine kinase; CSA: cross-sectional area; FFM: fat free mass; FOXO3: forkhead box O3; FSH: follicle-stimulating hormone; HRT: hormone replacement therapy; IGF: insulin-like growth factor; IGFBP: insulin-like growth factor binding protein; IL-6: interleukin-6; ILK: integrin linked kinase; LDH: lactate dehydrogenase; MVC: maximum voluntary contraction; RM: repetition maximum; STRAW: Stages of Reproductive Aging Workshop (Harlow et al., 2012); TCA cycle: the citric acid cycle. N/A = not applicable.

#### Muscle function

Nine studies investigated muscle function across different menopausal stages. Handgrip strength was significantly lower in post-versus premenopausal females in three studies (Preisinger *et al*., 1995; Laakkonen *et al*., 2017; Rathnayake *et al*., 2021) but remained similar in one study that compared age-matched pre-, peri- and postmenopausal females (Bassey *et al*., 1996). Postmenopausal females had weaker lower-limb strength than premenopausal females in some studies (Pöllänen *et al*., 2011; Pöllänen *et al*., 2015; Romero-Parra *et al*., 2021), but not in others (Buckley-Bleiler *et al*., 1989; Bassey *et al*., 1996; Laakkonen *et al*., 2017). Muscle power (as determined by vertical jump height or leg extensor power) was lower in post-versus premenopausal females in two studies (Pöllänen *et al*., 2015; Laakkonen *et al*., 2017), but again, this finding was not replicated in all (Bassey *et al*., 1996; Romero-Parra *et al*., 2021). Finally, when comparing 25 early to 38 late perimenopausal females, no differences in muscle function were reported, except for significantly lower *plantar flexor* strength in late perimenopausal females (Pesonen *et al*., 2021).

#### Muscle damage and repair

Five studies investigated the effect of menopausal status on the skeletal muscle damage and repair response. Cellular membrane damage leads to leakage of cytoplasmic molecules into circulation, such as creatine kinase (CK), lactate dehydrogenase (LDH) and myoglobin, making them indirect indicators of muscle damage after an exercise bout or injury (Clarkson & Hubal, 2002). Arnett *et al*. (2000) found that baseline serum CK levels were significantly greater in 10 post-versus 15 premenopausal females, and that serum CK was positively associated with serum E2. In contrast, after a muscle damage-inducing exercise bout, serum CK levels were lower in postmenopausal females compared to premenopausal females. Others, however, reported no such differences at rest or after exercise (Buckley-Bleiler *et al*., 1989; Romero-Parra *et al*., 2021). Nineteen postmenopausal females had significantly higher baseline levels of serum myoglobin compared to 13 premenopausal females prior to a damage-inducing bout of exercise, however serum LDH and muscle soreness were similar between groups before and after the intervention (Romero-Parra *et al*., 2021). Muscle-specific gene expression of pro-inflammatory cytokine IL-6 was similar between pre-(N=14) and postmenopausal (N=15) groups (Ahtiainen *et al*., 2012). Collins *et al*. (2019) found no difference in satellite cell number between peri- and postmenopausal females but did report a positive correlation between serum E2 levels and satellite cell number in both groups, although the sample size was very small (N=5 per group).

#### Energy metabolism

Two studies reported muscle outcomes relating to mitochondria and substrate metabolism and their association with menstrual status. Proteomic analysis by Laakkonen *et al*. (2017) found differential expression of pathways involved in energy metabolism, including mitochondrial dysfunction, oxidative phosphorylation, glycolysis and the citric-acid cycle between pre- and postmenopausal females in a sample size of 6 and 10 per group, respectively. In contrast, Nyberg *et al*. (2017) found no differences in the muscle protein content of mitochondrial complexes I to V or lactate release into the bloodstream of 20 pre- and 16 and early postmenopausal females.

Overall, these studies generally report reduced skeletal muscle mass and function in post-versus premenopausal females, suggesting a possible association with E2 decline. Evidence regarding the effect of menopause on the muscle damage response and energy metabolism however warrants further investigations.

### Effect of oestrogen supplementation on skeletal muscle health in postmenopausal females

#### Muscle mass

Eleven studies investigated differences in skeletal muscle outcomes depending on oestrogen supplementation status (Table 3). Of these, three investigated muscle mass outcomes. Taaffe *et al*. (2005) found that thigh muscle CSA was greater in a large cohort of 180 oestrogen replacement therapy (ORT) users versus 581 non-users, while Ryan *et al*. (2002) reported no differences in an overweight/obese population of only 6 participants per group. Interestingly, a positive association between the age of starting ORT and ALM was found in a cohort of 189 postmenopausal females (Kenny *et al*., 2003). As the dose and duration of ORT varied between these three studies, or was not specified, the conclusions that can be drawn remain limited.

**Table 3:** Studies investigating the effect of oestrogen supplementation on skeletal muscle outcomes in postmenopausal females.

| Study | Participants (mean age ± SD) | Oestrogen Use/Intervention | Exercise Intervention | Relevant Outcome Measures | Key Findings |
| --- | --- | --- | --- | --- | --- |
| *Cauley et al. (1987)* | 255 ORT users ($57 \pm 5$ years)<br>55 ORT non-users ($58 \pm 4$ years) | ORT users were prescribed treatment for menopausal symptoms or osteoporosis prevention (daily oestrogen dose: $0.37 \pm 0.19$mg, duration: $7.6 \pm 5.4$ years) | N/A | Handgrip strength | ↑ handgrip strength in ORT users ($27.5 \pm 4.0$kg versus $25.3 \pm 4.7$kg)<br>Positive association between oestrogen dose and handgrip strength ($r=0.16$)<br>Inverse association between years since menopause and handgrip strength ($r=-0.13$) |
| *Hansen et al. (2012)* | 10 ORT users ($60 \pm 4$ years)<br>10 ORT non-users ($61 \pm 4$ years) | ORT users had been prescribed treatment of oral E2 (dose: 2mg/day) after hysterectomy/oophorectomy (duration: $16.1 \pm 3.0$ years) | Resistance exercise session consisting of 10 sets of 10 reps of one-legged knee extension at 10-RM | Leg extensor 5-RM, myofibrillar protein FSR | ↔ leg extensor 5-RM (ORT: $27 \pm 9$kg, non-user: $31 \pm 9$kg)<br>↓ myofibrillar protein FSR at rest in ORT-users<br>↔ myofibrillar protein FSR 24 hours post-exercise<br>↑ myofibrillar protein FSR in exercised leg versus control leg in ORT users only |
| *Kenny et al. (2003)* | 189 ORT users ($68 \pm 5$ years) | Unknown type of oestrogen (dose: unknown, duration: $> 2$ years) | N/A | N/A | Positive association between age that participants started ORT and ALM ($r=0.158$) |
| *Silverstein et al. (1994)* | 465 previous or current ORT users ($\geq 65$ years)<br>176 ORT non-users ($\geq 65$ years) | Participants were classified into those who had or had not ever used ORT (dose and duration: unknown) | N/A | Handgrip strength | ↔ handgrip strength |
| *Maddalozzo et al. (2004)* | 67 ORT users ($51 \pm 3$ years)<br>59 ORT non-users ($51 \pm 3$ years) | ORT was conjugated equine oestrogen (dose: 0.625mg, duration: unknown) | N/A | Total body strength (measured from quadriceps, hamstring, hip abductor, pectoral, and *latissimus dorsi* muscles) and lower-limb explosive power | ↔ change in total body strength after 12 months (ORT: +9.8%, non-user: +10.8%)<br>↔ change in lower-limb explosive power after 12 months (ORT: +15.1%, non-user: +13.1%) |
| *Park et al. (2017) and Park et al. (2019)\** | 13 early postmenopausal females ($\leq 6$ years post-menopause, $55 \pm 3$ years)<br>14 late postmenopausal females ($\geq 10$ years post-menopause, $62 \pm 3$ years) | Random allocation to 1 week of transdermal E2 patch (dose: 0.15mg/week) or placebo patch, followed by 6-week wash-out period and other treatment | N/A | Muscle protein expression (Akt, pAkt, FOXO3, pFOXO3, MuRF1, and oxidative phosphorylation complexes II, IV, and V), nuclear and cytosolic content of PGC-1α, AMPK phosphorylation | ↔ Akt, pAkt, FOXO3, and pFOXO3 expression between groups<br>Group x treatment effect on FOXO3 dephosphorylation:<br>↑FOXO3 dephosphorylation (+68%) in late postmenopausal group following E2<br>↓FOXO3 dephosphorylation (-20%) in early postmenopausal group following E2<br>Group x treatment effect on MuRF1 expression:<br>↑MuRF1 expression (+17%) in late postmenopausal group following E2<br>↓ MuRF1 expression (-31%) early postmenopausal group following E2<br>↔ protein expression of mitochondrial complexes II, IV, and V between groups<br>↑PGC-1α nuclear protein content (+22%) in early postmenopausal group following E2<br>↓PGC-1α nuclear protein content (-23%) in late postmenopausal group following E2<br>↑ AMPK phosphorylation (-14%) in early postmenopausal group following E2<br>↓AMPK phosphorylation (-10%) in late postmenopausal group following E2 |
| *Pingel et al. (2012)* | 11 females ($65 \pm 2$ years) | Transdermal E2 patches for 5 days (dose: 2 x 50μg/day patches) or control period followed by 30 days of washout and the other treatment | Exercise session on day 2 of ORT: 10 reps of knee extension at 10-RM | Muscle IL-6 and IGF-1 | ↔ muscle IL-6 before and after exercise session<br>↔ muscle IGF-1 before and after exercise session |
| *Ryan et al. (2002)* | 6 overweight/obese ORT users ($57 \pm 3$ years)<br>6 overweight/obese ORT non-users ($58 \pm 2$ years) | ORT users took either conjugated oestrogen (dose: 0.625mg/day), or E2 (dose: 1mg/day or 0.5mg/day, duration: >3 years prior to study) | N/A | Thigh muscle CSA | ↔ thigh muscle CSA in ORT users versus non-users ($85.8 \pm 9.3\text{cm}^2$ versus $63.3 \pm 7.9\text{cm}^2$) |
| *Smith et al. (2014)* | 12 females ($61 \pm 2$ years) | Participants were randomised to control group or to receive transdermal E2 for 14 days (0.1mg/day), repeated twice more with 14-day non-treatment periods in between | N/A | Basal protein synthesis rate, mRNA expression of MyoD, myostatin, follistatin, and FOXO3 | ↔ basal protein synthesis and MyoD, myostatin, follistatin and FOXO3 expression between ORT users and non-users |
| *Taaffe et al. (1995)* | 37 ORT users (68 ± 1 years)<br>48 ORT non-users (70 ± 1 years) | 36/37 ORT users took conjugated equine oestrogen or equivalent (dose: 0.625mg/day, duration: 2-43 years) | N/A | 1-RM for leg press, knee extension and flexion, and hip abduction and adduction | ↔ leg press, knee extension and flexion, hip adduction and abduction 1-RM<br>No association between years of ORT use and muscle strength |
| *Taaffe et al. (2005)* | 180 current ORT users (range: 70-79 years)<br>581 current ORT non-users (74 ± 3 years) | Unspecified oestrogen-only hormone replacement therapy (dose and duration: unknown) | N/A | Quadriceps and hamstrings muscle CSA, quadriceps and hamstrings attenuation, handgrip strength, knee extensor torque and specific torque | ↑ quadriceps muscle CSA in ORT users (40.2 ± 0.4cm<sup>2</sup> versus 39.0 ± 0.2cm<sup>2</sup>)<br>↔ hamstring muscle CSA (users: 21.1 ± 0.3cm<sup>2</sup>, non-users: 21.3 ± 0.1cm<sup>2</sup>)<br>↔ quadriceps attenuation (users: 40.3 ± 0.4HU, non-users: 39.4 ± 0.2HU)<br>↔ hamstring attenuation (users: 28.0 ± 0.6HU, non-users: 27.4 ± 0.3HU)<br>↔ handgrip strength (users: 45.6 ± 0.7kg, non-users: 44.1 ± 0.4kg)<br>↔ knee extensor torque (users: 79.3 ± 1.4kg, non-users: 77.7 ± 0.8kg)<br>↔ knee extensor specific torque (users: 1.95 ± 0.03N/m/cm<sup>2</sup>, non-users: 1.98 ± 0.02N/m/cm<sup>2</sup>) |

↑ = significant increase, ↓ = significant decrease, ↔ no difference, $p < 0.05$ . Akt: protein kinase B; ALM: appendicular lean mass; AMPK: adenosine monophosphate-activated protein kinase; CSA: cross-sectional area; FOXO3: forkhead box O3; FSR: fractional synthesis rate; IGF-1: insulin-like growth factor 1; IL-6: interleukin 6; MHC: myosin heavy chain; MuRF1: muscle ring-finger protein 1; ORT: oestrogen replacement therapy; p-Akt: phosphorylated protein kinase B; PGC-1α: peroxisome proliferator-activated receptor γ co-activator 1α; p-FOXO3: phosphorylated forkhead box O3; RCT: randomised controlled trial; RM: repetition maximum; N/A = not applicable

Three studies further explored the effects of ORT on the molecular regulation of muscle mass. In 10 long-term ORT users (duration: 16.1 ± 3.0 years), myofibrillar protein fractional synthesis rate was lower than in 10 non-users at rest and after a bout of resistance exercise (Hansen *et al*., 2012). Protein content of the total and phosphorylated form of the muscle protein synthesis marker protein kinase B (Akt), baseline protein fractional synthesis rate and mRNA expression of catabolic markers myostatin and FOXO3 were similar after short-term (1 week (Park *et al*., 2019) to 6 weeks (Smith *et al*., 2014)) ORT use versus non-users. Finally, Park *et al*. (2019) found that one week of E2 supplementation increased the protein expression of muscle-ring finger protein 1 (MuRF1) and decreased the phosphorylation status of FOXO3 in 14 late postmenopausal females, but had the opposite effect in 13 early postmenopausal females.

#### Muscle function

Six studies investigated the effect of ORT on skeletal muscle function. One study found that handgrip strength was greater in a cohort of 255 long-term ORT users (duration: 7.6 ± 5.4 years) versus 55 non-users, and was positively associated with E2 dose (Cauley *et al*., 1987), but this was not replicated by other large studies (Kritz-Silverstein & Barrett-Connor, 1994; Taaffe *et al*., 2005). It should be noted however these studies did not report participants’ dose or duration of ORT. There was no effect of long-term ORT on lower-limb strength and power (Taaffe *et al*., 1995; Taaffe *et al*., 2005; Hansen *et al*., 2012) and total body strength and lower-limb power did not differ between users and non-users in early menopause (<36 months) or after a 12-month follow up (Maddalozzo *et al*., 2004).

#### Energy metabolism

One acute study examined the effect of ORT in mechanisms of energy metabolism. Following one week of E2 supplementation, adenosine monophosphate-activated protein kinase (AMPK) phosphorylation and the nuclear protein content of muscle peroxisome proliferator-activated receptor γ co-activator 1α (PGC-1α) were increased in 13 early postmenopausal females, but decreased in 14 late postmenopausal females (Park *et al*., 2017). Protein expression of the mitochondrial complexes II, IV, and V was not different between groups (Park *et al*., 2017).

#### Muscle damage and repair

Finally, one cross-over study investigated the effect of five days or E2 supplementation on muscle inflammation and found no differences of treatment on muscle IL-6 content in 11 postmenopausal participants, both before and after a resistance exercise bout (Pingel *et al*., 2012).

Overall, the literature regarding the effect of oestrogen supplementation on the composition and molecular regulation of muscle mass, muscle strength, energy metabolism and inflammation does not provide any conclusive evidence.

## Discussion

### Effect of oestrogen on ageing skeletal muscle mass

Overall, postmenopausal females had lower muscle mass and CSA than premenopausal females (Pöllänen *et al*., 2011; Pöllänen *et al*., 2015; Nyberg *et al*., 2017; Juppi *et al*., 2020; Park *et al*., 2020; Rathnayake *et al*., 2021). However, in postmenopausal females, the relationship between oestrogen status and muscle mass was unclear in both ORT users (Ryan *et al*., 2002; Taaffe *et al*., 2005) and non-users (Kenny *et al*., 2003; Rolland *et al*., 2007; García-Martín *et al*., 2013; Gonnelli *et al*., 2014; Kong *et al*., 2019; Guligowska *et al*., 2021). This suggests that the reduction in E2 may be a factor in the menopause-induced loss of muscle mass, but that its role may diminish after the menopausal transition. Studies comparing pre- and postmenopausal females provide a useful model to investigate the effect of E2 deficiency, but singling out the effects of menopause from other age-related factors is challenging. ORT studies are therefore beneficial to further understand the specific role of oestrogen without the confounding effect of age. The large variability found in ORT type, dose and duration is however a strong limitation to the interpretation of these findings. Animal models generally demonstrate that sex hormone deficiency (induced by ovariectomy, (OVX)) reduces muscle mass and that E2 treatment may partially reverse these negative effects (Pellegrino *et al*., 2022).

No human studies examined differences in the molecular regulation of muscle mass between pre- and postmenopausal females, but a few investigated the effect of ORT. Of these, there were minimal differences in anabolic and catabolic signalling markers, suggesting exogenous oestrogen supplementation does not significantly impact the molecular signalling of protein turnover in the short term (Pingel *et al*., 2012; Smith *et al*., 2014; Park *et al*., 2019). In contrast, as reviewed in Pellegrino *et al*. (2022), animal studies have shown that OVX increases the expression of catabolic factors (e.g. MuRF1and FOXO3) and decreases the expression of anabolic factors (e.g. phosphoinositide 3 kinase (PI3K) and Akt). These effects can be reversed by E2 treatment, suggesting a direct or indirect role of oestrogen in the molecular regulation of muscle protein turnover. Whilst oestrogen deficiency seems to elicit a negative protein balance that potentially contributes to the resulting loss of muscle mass in animal model, this pattern is not replicated in humans. This may be explained by the less extreme nature of human oestrogen deficiency models (menopause) when comparted to animal (OVX) studies. It may however be too simplistic to argue that oestrogen has a purely anabolic role within the muscle. Indeed, Hansen *et al*. (2012) found lower basal protein fractional synthesis rates in a small study comprising 10 long-term ORT users compared to 10 non-users, a pattern that has also been shown in rats (Toth *et al*. (2001). Rat studies suggest that oestrogen may also contribute to protein breakdown by activating the ubiquitin proteasome system (Beckett *et al*., 2002) and autophagy (Zhong *et al*., 2019) pathways. The role of oestrogen in protein turnover also appears to be dependent on internal factors, such as when the individual becomes sex-hormone deficient. For example, a small human study (Park *et al*., 2020) suggested that the time since menopause may impact ORT-induced changes in protein breakdown factors, MuRF1 and FOXO3, as opposing effects were found in early versus late postmenopausal females. The age that individuals started ORT was also correlated with muscle mass in a large participant cohort (Kenny *et al*., 2003). This idea is supported by animal studies, where both Mangan *et al*. (2015) and (Tsai *et al*., 2007) found differential effects of oestrogen treatment dependent on timing of treatment after OVX.

More high-quality and well-controlled human research is needed to fully elucidate the role of oestrogen on muscle mass throughout ageing and how its levels may be manipulated for better health outcomes.

### Effect of oestrogen on ageing skeletal muscle function

No correlations were found between serum E2 and muscle function after menopause (Bochud *et al*., 2019; Kong *et al*., 2019; Guligowska *et al*., 2021). In the only study that examined the role of E1, the predominant type of oestrogen after menopause, E1 was positively associated with postmenopausal loss of muscle strength (Rolland *et al*., 2007). Changes in muscle strength across the menopausal transition were generally incongruent between studies. Most indicated a reduction in handgrip strength after menopause (Preisinger *et al*., 1995; Laakkonen *et al*., 2017; Rathnayake *et al*., 2021), which is a reliable indicator of whole-body strength (Trosclair *et al*., 2011), but changes in lower-limb strength and power were mixed. Evidence of the effect of ORT on muscle function was also limited. Long-term ORT users had greater handgrip strength in one study (Cauley *et al*., 1987), but not others (Kritz-Silverstein & Barrett-Connor, 1994; Taaffe *et al*., 2005), and there was no effect of ORT use on changes in lower-limb strength and power after 12-months (Maddalozzo *et al*., 2004).

The large variability in ORT type, dose, and duration is again a major limitation to these findings. Only one study found increased muscle function with long-term users of ORT (mean duration: 7.6 years ± 5.4 years). While three other studies showed no effect of ORT, the duration of the treatment was not stated, and one of these included previous (not current) users in their ORT group, making it difficult to determine the true effect of oestrogen supplementation of muscle function. In contrast, in animal models, oestrogen deficiency clearly contributes to a loss of grip strength and force production, the effects of which can be reversed by E2 treatment (reviewed in Pellegrino *et al*., 2022).

### Limitations

The general lack of clear evidence regarding the effect of exogenous oestrogen supplementation can be partly explained by the high variability associated with the doses and durations of oestrogen treatment. In three of the ORT studies, supplementation ranged from 5-14 days, which may not have been enough time for oestrogen to exert its potential effects on muscle protein turnover, while the effect of long-term oestrogen supplementation on the molecular regulation of muscle mass has not been investigated at all. There was also large variation and a lack of reporting of ORT type, dose, and duration, which can be partly explained by the inclusion of case-control studies. Some studies investigated long-term ORT users (> 2 years, n=4), while other treatment programs were short-term (< 3 months, n=4), and some studies did not report this information at all (n=3).

Additional limitations to this review include notable variation in the criteria used to define menopausal status. Of the 17 studies that investigated muscle differences between menopausal stages, 10 did not report the criteria they used to define the stages. Five studies used a combination of menstrual cycle tracking (e.g. date of last known menstrual bleed) and serum hormone analysis to determine the menopausal stage, and only three of these stated the defining criteria. Similarly, of the 13 studies that included premenopausal females, only one specified if and how they measured and controlled for menstrual cycle phase. In premenopausal females, some studies have shown that menstrual cycle phase can influence skeletal muscle function and metabolism (Knowles *et al*., 2019), highlighting the need for more tightly-controlled studies.

Variability in the methodology used to assess muscular outcomes may have further contributed to the incongruence across studies. For example, there were differing correlations between E2 and muscle when assessed by BIA (Guligowska *et al*., 2021) or DXA (García-Martín *et al*., 2013; Gonnelli *et al*., 2014). There was also large variation in the method (repetition-max, dynamometry) and location (e.g., hand, leg, finger, foot, and total body) used to assess muscle strength, adding an extra layer of complexity when comparing findings from different studies.

Finally, 40% of studies had a moderate or high risk of bias. While confounding variables were identified in most (73%) cross-sectional studies, 50% did not state whether these were statistically adjusted for (Supplementary File 5). As a result, in some cases, it could not be confirmed whether the observed differences were due to menopausal or ORT user status. Further, variability in diet, physical activity levels, or training status may also confound results as they all have the potential to influence skeletal muscle metabolism. Lastly, none of the RCTs described their randomisation procedure, or if participants and assessors had been blinded to the assigned conditions (Supplementary File 5).

Together, these limitations partly explain why our human findings were largely conflicting and limited conclusions could be made. Future research in the field must be more transparent about measurements of the menstrual cycle and/or menopausal status, where relevant, and exogenous oestrogen supplementation to allow suitable application and generalisation of the findings.

### Conclusion

While the findings of this systematic review were largely inconsistent, there is some evidence to suggest that oestrogen may have a beneficial role in the maintenance of skeletal muscle during female ageing. Studies have demonstrated that sex hormone deficiency is associated with poorer outcomes for measures of muscle mass, function, damage and repair and energy metabolism, some of which could potentially be improved by exogenous oestrogen supplementation. However, the overall quality of the literature is a limitation. There is a strong need for higher quality human evidence to fully elucidate how the decline in endogenous oestrogen production with ageing impacts female skeletal muscle. This will allow further understanding of female-specific mechanisms of muscle ageing. Only high-quality literature will allow the identification and implementation of appropriate strategies to prevent and manage age-related functional decline within the growing population of older females.

## Data Availability

All data produced in the present work are contained in the manuscript and supplementary files.

## Additional Information

### Author contributions

AC, DH, DS, and SL contributed to the conception and design of the review. AC and RW acquired, analysed, and interpreted the data. AC and SL drafted the review, and DH, DS, and RW provided critical revisions.

## Supporting Information

- Supplementary File 1 – PRISMA 2020 Checklist
- Supplementary File 2 – PRISMA Abstract Checklist
- Supplementary File 3 - AMSTAR2 Tool
- Supplementary File 4 - Systematic Review Methods
- Supplementary File 5 – Quality Assessment Results Tables

