## Supplementary file 3 for "The Role of Oestrogen in Female Skeletal Muscle Ageing: A Systematic Review"

### AMSTAR 2: a critical appraisal tool for systematic reviews that include randomised or non-randomised studies of healthcare interventions, or both

**1. Did the research questions and inclusion criteria for the review include the components of PICO?**

For Yes:
- ☒ Population
- ☒ Intervention
- ☒ Comparator group
- ☒ Outcome

Optional (recommended)
- ☐ Timeframe for follow-up

- ☒ Yes
- ☐ No

**2. Did the report of the review contain an explicit statement that the review methods were established prior to the conduct of the review and did the report justify any significant deviations from the protocol?**

For Partial Yes:
The authors state that they had a written protocol or guide that included ALL the following:
- ☒ review question(s)
- ☒ a search strategy
- ☒ inclusion/exclusion criteria
- ☒ a risk of bias assessment

For Yes:
As for partial yes, plus the protocol should be registered and should also have specified:
- ☐ a meta-analysis/synthesis plan, if appropriate, *and*
- ☒ a plan for investigating causes of heterogeneity
- ☒ justification for any deviations from the protocol

- ☒ Yes
- ☐ Partial Yes
- ☐ No

**3. Did the review authors explain their selection of the study designs for inclusion in the review?**

For Yes, the review should satisfy ONE of the following:
- ☐ *Explanation for* including only RCTs
- ☐ OR *Explanation for* including only NRSI
- ☐ OR *Explanation for* including both RCTs and NRSI

- ☐ Yes
- ☒ No

**4. Did the review authors use a comprehensive literature search strategy?**

For Partial Yes (all the following):
- ☒ searched at least 2 databases (relevant to research question)
- ☒ provided key word and/or search strategy
- ☐ justified publication restrictions (e.g. language)

For Yes, should also have (all the following):
- ☐ searched the reference lists / bibliographies of included studies
- ☐ searched trial/study registries
- ☐ included/consulted content experts in the field
- ☐ where relevant, searched for grey literature
- ☒ conducted search within 24 months of completion of the review

- ☐ Yes
- ☒ Partial Yes
- ☐ No

**5. Did the review authors perform study selection in duplicate?**

For Yes, either ONE of the following:
- ☒ at least two reviewers independently agreed on selection of eligible studies and achieved consensus on which studies to include
- ☐ OR two reviewers selected a sample of eligible studies and achieved good agreement (at least 80 percent), with the remainder selected by one reviewer.

- ☒ Yes
- ☐ No

##### 6. Did the review authors perform data extraction in duplicate?

| | |
|---|---|
| For Yes, either ONE of the following:<br>☒ at least two reviewers achieved consensus on which data to extract from included studies<br>☐ OR two reviewers extracted data from a sample of eligible studies <u>and</u> achieved good agreement (at least 80 percent), with the remainder extracted by one reviewer. | ☒ Yes<br>☐ No |

##### 7. Did the review authors provide a list of excluded studies and justify the exclusions?

| | | |
|---|---|---|
| For Partial Yes:<br>☐ provided a list of all potentially relevant studies that were read in full-text form but excluded from the review | For Yes, must also have:<br>☒ Justified the exclusion from the review of each potentially relevant study | ☐ Yes<br>☒ Partial Yes<br>☐ No |

##### 8. Did the review authors describe the included studies in adequate detail?

| | | |
|---|---|---|
| For Partial Yes (ALL the following):<br>☒ described populations<br>☒ described interventions<br>☒ described comparators<br>☒ described outcomes<br>☒ described research designs | For Yes, should also have ALL the following:<br>☒ described population in detail<br>☒ described intervention in detail (including doses where relevant)<br>☒ described comparator in detail (including doses where relevant)<br>☒ described study's setting<br>☒ timeframe for follow-up | ☒ Yes<br>☐ Partial Yes<br>☐ No |

##### 9. Did the review authors use a satisfactory technique for assessing the risk of bias (RoB) in individual studies that were included in the review?

| | | |
|---|---|---|
| **RCTs**<br>For Partial Yes, must have assessed RoB from<br>☒ unconcealed allocation, *and*<br>☒ lack of blinding of patients and assessors when assessing outcomes (unnecessary for objective outcomes such as all-cause mortality) | For Yes, must also have assessed RoB from:<br>☒ allocation sequence that was not truly random, *and*<br>☒ selection of the reported result from among multiple measurements or analyses of a specified outcome | ☒ Yes<br>☐ Partial Yes<br>☐ No<br>☐ Includes only NRSI |
| **NRSI**<br>For Partial Yes, must have assessed RoB:<br>☒ from confounding, *and*<br>☒ from selection bias | For Yes, must also have assessed RoB:<br>☒ methods used to ascertain exposures and outcomes, *and*<br>☒ selection of the reported result from among multiple measurements or analyses of a specified outcome | ☒ Yes<br>☐ Partial Yes<br>☐ No<br>☐ Includes only RCTs |

##### 10. Did the review authors report on the sources of funding for the studies included in the review?

| | |
|---|---|
| For Yes<br>☐ Must have reported on the sources of funding for individual studies included in the review. Note: Reporting that the reviewers looked for this information but it was not reported by study authors also qualifies | ☐ Yes<br>☒ No |

##### 11. If meta-analysis was performed did the review authors use appropriate methods for statistical combination of results?

**RCTs**
For Yes:

- ☐ The authors justified combining the data in a meta-analysis
  - ☐ AND they used an appropriate weighted technique to combine study results and adjusted for heterogeneity if present.
  - ☐ AND investigated the causes of any heterogeneity

☐ Yes
☐ No
☒ No meta-analysis conducted

**For NRSI**
For Yes:

- ☐ The authors justified combining the data in a meta-analysis
  - ☐ AND they used an appropriate weighted technique to combine study results, adjusting for heterogeneity if present
  - ☐ AND they statistically combined effect estimates from NRSI that were adjusted for confounding, rather than combining raw data, or justified combining raw data when adjusted effect estimates were not available
  - ☐ AND they reported separate summary estimates for RCTs and NRSI separately when both were included in the review

☐ Yes
☐ No
☒ No meta-analysis conducted

##### 12. If meta-analysis was performed, did the review authors assess the potential impact of RoB in individual studies on the results of the meta-analysis or other evidence synthesis?

For Yes:

- ☐ included only low risk of bias RCTs
- ☐ OR, if the pooled estimate was based on RCTs and/or NRSI at variable RoB, the authors performed analyses to investigate possible impact of RoB on summary estimates of effect.

☐ Yes
☐ No
☒ No meta-analysis conducted

##### 13. Did the review authors account for RoB in individual studies when interpreting/ discussing the results of the review?

For Yes:

- ☐ included only low risk of bias RCTs
- ☒ OR, if RCTs with moderate or high RoB, or NRSI were included the review provided a discussion of the likely impact of RoB on the results

☒ Yes
☐ No

##### 14. Did the review authors provide a satisfactory explanation for, and discussion of, any heterogeneity observed in the results of the review?

For Yes:

- ☐ There was no significant heterogeneity in the results
- ☒ OR if heterogeneity was present the authors performed an investigation of sources of any heterogeneity in the results and discussed the impact of this on the results of the review

☒ Yes
☐ No

##### 15. If they performed quantitative synthesis did the review authors carry out an adequate investigation of publication bias (small study bias) and discuss its likely impact on the results of the review?

For Yes:

- ☐ performed graphical or statistical tests for publication bias and discussed the likelihood and magnitude of impact of publication bias

☐ Yes
☐ No
☒ No meta-analysis conducted

**16. Did the review authors report any potential sources of conflict of interest, including any funding they received for conducting the review?**

For Yes:

| | | | |
|---|---|---|---|
| ☒ | The authors reported no competing interests OR | ☒ | Yes |
| ☐ | The authors described their funding sources and how they managed potential conflicts of interest | ☐ | No |

**To cite this tool:** Shea BJ, Reeves BC, Wells G, Thuku M, Hamel C, Moran J, Moher D, Tugwell P, Welch V, Kristjansson E, Henry DA. AMSTAR 2: a critical appraisal tool for systematic reviews that include randomised or non-randomised studies of healthcare interventions, or both. BMJ. 2017 Sep 21;358:j4008.
