## Supplementary file 4 for "The Role of Oestrogen in Female Skeletal Muscle Ageing: A Systematic Review"

***Supplementary File 3 - Methods***

*Database Searching*

A systematic search was carried out using 6 electronic databases, including MEDLINE complete, Global Health, Embase, PubMed, SPORTDiscus, and CINHAL. Search restrictions included journal articles in the English language, and search terms in the title or abstract fields. Key word search terms were 1) oestrogen* OR estrogen* OR oestradiol OR estradiol OR oestrone OR estrone OR oestriol OR estriol OR “sex hormone*” OR “sex steroid*”, 2) female* OR wom?n, 3) muscle* OR “skeletal muscle*”, and 4) ageing OR “old* adult*” OR elderly OR lifespan OR maturing OR senior OR geriatric* OR postmenopaus* OR menopaus* OR “post menopaus*” OR ovariectom*. These 4 terms were combined with ‘AND’. Databases were searched from inception to 08/11/2022.

*Eligibility Criteria*

The Population, Intervention, Comparison, Outcomes and Study design (PICOS) framework was used to define the study eligibility criteria. The eligibility criteria required studies to investigate the role of oestrogen in skeletal muscle health in a model of female ageing. Inclusion criteria consisted of quantitative human studies that analysed a model of oestrogen deficiency (e.g. postmenopausal females) or supplementation (e.g. oestrogen treatment, hormonal replacement therapy (HRT), or hormonal contraception), and compared it to normal oestrogen conditions (e.g. premenopausal females, no supplementation). Longitudinal, cross-sectional, observational, and experimental studies were included. Outcome variables of interest were compositional, functional, or molecular measures of skeletal muscle mass, strength, damage, regeneration, inflammation, and energy metabolism. Exclusion criteria included qualitative studies, training intervention studies with no relevant baseline measures, no female-specific analysis, the presence of a chronic disease without healthy comparisons, no report of hormonal status or HRT/hormonal contraception type, use of combined oestrogen and progesterone treatment, and no relevant skeletal muscle health outcomes. Total lean body mass was excluded as a measure of muscle mass, as this also reflects residual soft tissue such as organs, particularly in the abdomen region (Evans et al., 2019). To minimise this inaccuracy, only measures of appendicular lean mass (sum of arms and legs) were included, as is commonly used in sarcopenia research (Cawthorn, 2015).

*Screening and Quality Assessment*

Using Covidence systematic review software (Veritas Health Innovation, Melbourne, Australia), duplicate articles were removed, followed by a 2-phase screening strategy. First, titles and abstracts were screened for eligibility, and second, full text screening was conducted to remove any studies that did not meet before the eligibility criteria. The quality of the included studies was assessed using The Johanna Briggs Institute critical appraisal tool for the relevant study design (cross-sectional, quasi-experimental, randomised controlled trials (RCTs), and cohort) (Moola et al. 2020). Risk of bias was calculated by the percentage of ‘yes’ answers: 1) ≤49% = high risk, 2) 50-69% = moderate risk, and 3) ≥70% = low risk. Screening and quality assessment was carried out by 2 reviewers (AC, RW). If disagreements occurred, it was first discussed between reviewers, and if remained unsolved, it was resolved by a third reviewer (DH).

*Data Extraction and Synthesis*

Data were extracted from all eligible studies independently by two reviewers (AC and RW) using an *a-priori* designed data extraction form. Data extracted included author information, participant characteristics, study design, methodology, findings, and study limitations. Data regarding irrelevant outcome measures were not extracted, decided by pre-specified outcome measures of interest. Synthesised data were based only on data available in the manuscript or supplementary materials. If disagreements occurred, it was first discussed between reviewers (AC, RW), and if remained unsolved, it was resolved by a third reviewer (DH). After data extraction, the findings of the systematic review were summarised according to the study design in a narrative review. Studies designs included studies investigating associations between oestrogen levels and muscle outcomes, studies investigating differences in skeletal muscle outcomes depending on menopausal status and studies investigating differences in skeletal muscle outcomes depending on oestrogen supplementation status. Where studies fit into more than one category, specific outcomes were reported in the relevant categories. Within these three study designs, findings were reported on the following outcome measures: skeletal muscle mass and composition, skeletal muscle function, skeletal muscle damage and repair, and energy metabolism. Due to the broad range of study designs included in the search, there was no quantitative meta-analysis of the data.

*Literature Selection*

Based on the database search, 4059 articles were retrieved and imported into Covidence for screening. A total of 1729 duplicates were removed, leaving 2330 references for title and abstract screening. Following title and abstract screening, 2025 articles were excluded based on the PICO criteria, leaving 305 articles identified for the full text screen. Based on the full text screen, 272 studies were excluded for the following reasons: animal studies; used combined oestrogen and progesterone supplementation; no suitable outcome measures; no report of hormone supplementation type; training intervention studies; not original articles; unhealthy population; no comparison between specified groups; no female-specific analysis; full text unavailable; not in English. As two eligible papers were derived from the same study, they were classified as one study. In total 32 studies were included in the review.
