## Supplementary file 5 for "The Role of Oestrogen in Female Skeletal Muscle Ageing: A Systematic Review"

***Supplementary File 4 – Quality Assessment***

**Supplementary Table 1: Quality assessment of cross-sectional studies**

| **Study** | **Q1** | **Q2** | **Q3** | **Q4** | **Q5** | **Q6** | **Q7** | **Q8** | **% Yes** | **Risk of Bias** |
| --- | --- | --- | --- | --- | --- | --- | --- | --- | --- | --- |
| *Ahtianen et al. (2012)* | N | N | ? | ? | N | N | Y | Y | 25 | High |
| *Bassey et al. (1996)* | Y | N | N | N | ? | N | N | N | 12.5 | High |
| *Bochud et al. (2019)* | Y | Y | N | N | Y | Y | Y | Y | 75 | Low |
| *Cauley et al. (1987)* | Y | Y | Y | Y | Y | Y | Y | Y | 100 | Low |
| *Garcia-Martin et al. (2013)* | Y | Y | Y | Y | N | Y | Y | Y | 87.5 | Low |
| *Gonelli et al. (2014)* | Y | Y | Y | Y | Y | Y | Y | Y | 100 | Low |
| *Guligowska et al. (2021)* | Y | Y | Y | Y | N | ? | Y | Y | 75 | Low |
| *Hansen et al. (2012)* | Y | Y | Y | Y | Y | N | Y | Y | 87.5 | Low |
| *Kenny et al. (2003)* | Y | N | Y | N | Y | N | Y | N | 50 | Moderate |
| *Kong et al. (2019)* | N | N | Y | Y | Y | N | Y | Y | 62.5 | Moderate |
| *Kritz-Silverstein et al. (1994)* | Y | Y | N | Y | Y | Y | Y | Y | 87.5 | Low |
| *Lakkonen et al. (2017)* | N | Y | N | N | Y | N | Y | Y | 50 | Moderate |
| *Park et al. (2020)* | Y | Y | Y | Y | Y | Y | Y | Y | 100 | Low |
| *Personen et al. (2021)* | Y | Y | Y | Y | Y | N | Y | Y | 87.5 | Low |
| *Pollanen et al. (2011)* | Y | Y | N | N | Y | N | Y | Y | 62.5 | Moderate |
| *Pollanen et al. (2015)* | N | Y | N | ? | ? | Y | Y | Y | 50 | Moderate |
| *Preisinger et al. (1995)* | Y | Y | N | N | Y | Y | Y | N | 62.5 | Moderate |
| *Rathnayake et al. (2021)* | Y | Y | Y | Y | Y | Y | Y | Y | 100 | Low |
| *Romero-Parra et al. (2021)* | Y | Y | Y | Y | N | N | Y | Y | 75 | Low |
| *Ryan et al. (2002)* | Y | Y | N | N | Y | Y | Y | Y | 75 | Low |
| *Taaffe et al. (1995)* | Y | Y | ? | ? | Y | N | N | Y | 50 | Moderate |
| *Taaffe et al. (2005)* | Y | Y | ? | Y | Y | Y | Y | Y | 87.5 | Low |

*Q1: Were the criteria for inclusion in the sample clearly defined? Q2: Were the study subjects and the setting described in detail? Q3: Was the exposure measured in a valid and reliable way? Q4: Were objective, standard criteria used for measurement of the condition? Q5: Were confounding variables identified? Q6: Were strategies to deal with confounding variables stated? Q7: Were the outcomes measured in a vlid and reliable way? Q8: Was appropriate statistical analysis used? ≤49% yes = high risk of bias, 50-69% yes = moderate risk of bias, ≥70% yes = low risk of bias.*

**Supplementary Table 2: Quality assessment of quasi-experimental studies**

| **Study** | **Q1** | **Q2** | **Q3** | **Q4** | **Q5** | **Q6** | **Q7** | **Q8** | **Q9** | **% Yes** | **Risk of Bias** |
| --- | --- | --- | --- | --- | --- | --- | --- | --- | --- | --- | --- |
| *Arnett et al. (2000)* | Y | Y | ? | Y | Y | Y | Y | N | Y | 67 | Moderate |
| *Buckley-Bleiler et al. (1989)* | Y | Y | Y | Y | Y | Y | Y | ? | Y | 89 | Low |
| *Nyberg et al. (2017)* | Y | Y | Y | Y | Y | N | Y | Y | Y | 89 | Low |

*Q1: Is it clear in the study what is the 'cause' and what is the 'effect'? Q2: Were the participants included in any comparisons similar? Q3: Were the participants included in any comparisons receiving similar treatment/care, other than the exposure or intervention of interest? Q4: Was there a control group? Q5: Were there multiple measurements of the outcome both pre and post the intervention/exposure? Q6: Was follow up complete and if not, were differences between groups in terms of their follow up adequately described and analysed? Q7: Were the outcomes of participants included in any comparisons measured in the same way? Q8: Were outcomes measured in a reliable way? Q9: Was appropriate statistical analysis used? ≤49% yes = high risk of bias, 50-69% yes = moderate risk of bias, ≥70% yes = low risk of bias.*

**Supplementary Table 3: Quality assessment of randomised controlled trials**

| **Study** | **Q1** | **Q2** | **Q3** | **Q4** | **Q5** | **Q6** | **Q7** | **Q8** | **Q9** | **Q10** | **Q11** | **Q12** | **% Yes** | **Risk of Bias** |
| --- | --- | --- | --- | --- | --- | --- | --- | --- | --- | --- | --- | --- | --- | --- |
| *Park et al. (2017, 2019)* | ? | ? | Y | Y | ? | Y | Y | ? | Y | Y | Y | ? | 58 | Moderate |
| *Pingel et al. (2012)* | ? | ? | ? | ? | ? | Y | Y | ? | Y | ? | Y | N | 33 | High |
| *Smith et al. (2014)* | ? | ? | N | N | ? | Y | Y | ? | Y | Y | Y | Y | 50 | Moderate |

*Q1: Were the two groups similar and recruited from the same population? Q2: Were the exposures measured similarly to assign people to both exposed and unexposed groups? Q3: Was the exposure measured in a valid and reliable way? Q4: Were confounding factors identified? Q5: Were strategies to deal with confounding factors stated? Q6: Were the groups/participants free of the outcome at the start of the study (or at the moment of exposure)? Q7: Were the outcomes measured in a valid and reliable way? Q8: Was the follow up time reported and sufficient to be long enough for outcomes to occur? Q9: Was follow up complete, and if not, were the reasons to loss to follow up described and explored? Q10: Were strategies to address incomplete follow up utilized? Q11: Was appropriate statistical analysis used? ≤49% yes = high risk of bias, 50-69% yes = moderate risk of bias, ≥70% yes = low risk of bias.*

**Supplementary Table 4: Quality assessment of longitudinal studies**

| **Study** | **Q1** | **Q2** | **Q3** | **Q4** | **Q5** | **Q6** | **Q7** | **Q8** | **Q9** | **Q10** | **Q11** | **% Yes** | **Risk of Bias** |
| --- | --- | --- | --- | --- | --- | --- | --- | --- | --- | --- | --- | --- | --- |
| *Collins et al. (2019)* | Y | Y | Y | Y | Y | N/A | Y | N | Y | Y | Y | 90 | Low |
| *Juppi et al. (2020)* | Y | Y | Y | Y | Y | N/A | Y | ? | Y | Y | Y | 90 | Low |
| *Maddalozzo et al. (2004)* | Y | Y | Y | Y | Y | N/A | Y | Y | Y | Y | N | 90 | Low |
| *Rolland et al. (2007)* | Y | N/A | N | Y | Y | N/A | Y | Y | Y | N | Y | 78 | Low |

*Q1: Were the two groups similar and recruited from the same population? Q2: Were the exposures measured similarly to assign people to both exposed and unexposed groups? Q3: Was the exposure measured in a valid and reliable way? Q4: Were confounding factors identified? Q5: Were strategies to deal with confounding factors stated? Q6: Were the groups/participants free of the outcome at the start of the study (or at the moment of exposure)? Q7: Were the outcomes measured in a valid and reliable way? Q8: Was the follow up time reported and sufficient to be long enough for outcomes to occur? Q9: Was follow up complete, and if not, were the reasons to loss to follow up described and explored? Q10: Were strategies to address incomplete follow up utilized? Q11: Was appropriate statistical analysis used? ≤49% yes = high risk of bias, 50-69% yes = moderate risk of bias, ≥70% yes = low risk of bias.*
